## Supplementary material for "Higher ratio of plasma omega-6/omega-3 fatty acids is associated with greater risk of all-cause, cancer, and cardiovascular mortality: a population-based cohort study in UK Biobank": Table S1

**Table S1**. Literature Review

| Author | Year | Study design/ location | Endpoints | # Cohort | # Cases | Exact exposure | Exposure characterization | Measure of associations | P for trend |
|---|---|---|---|---|---|---|---|---|---|
| *Omega-6/omega-3 ratio* | | | | | | | | | |
| Noori et al.[1] | 2011 | Prospective cohort/ hemodialysis patients, Southern California during 2001-2007 | All-cause mortality | 145 | 42 | Dietary Omega-3<br>Dietary Omega-6/3 | HR in quartiles (95% CI) | (Highest to lowest) 0.65 (0.21-1.97)<br>(Lowest to highest) 0.37 (0.14-1.08) | 0.1<br>0.04 |
| Harris et al.[2] | 2017 | Prospective cohort/ women, Women's Health Initiative Memory Study, enrollment began 1996, age 65-80 | All-cause mortality<br><br><br><br><br><br><br><br>CVD mortality<br><br><br><br><br><br><br>Cancer mortality | 6,501 | 1,851<br><br><br><br><br><br><br><br>617<br><br><br><br><br><br><br>462 | Red blood cell Omega-3 index (EPA + DHA)<br>EPA<br>DHA<br>LA<br>N-6/N-3 ratio | HR per 1-SD increase in red blood cell PUFA (99% CI) | All<br>Omega-3 index: 0.92 (0.85-0.98)<br>EPA: 0.89 (0.82-0.96)<br>DHA: 0.93 (0.87-1.00)<br>LA: 0.99 (0.93-1.06)<br>N-6/N-3 ratio: 1.10 (1.02-1.19)<br>CVD<br>Omega-3 index: 0.97 (0.85-1.12)<br>EPA: 0.88 (0.77-1.00)<br>DHA: 1 (0.87-1.14)<br>LA: 0.96 (0.86-1.08)<br>N-6/N-3 ratio: 1.05 (0.9-1.23)<br>Cancer<br>Omega-3 index: 0.92 (0.79-1.07)<br>EPA: 0.91 (0.78-1.07)<br>DHA: 0.93 (0.79-1.09)<br>LA: 0.94 (0.83-1.07)<br>N-6/N-3 ratio: 1.1 (0.93-1.3) | <br>0.0015<br>< 0.001<br>0.0099<br>0.8286<br>0.0021<br>NR<br><br><br><br><br><br>NR |
| Otsuka et al.[3] | 2019 | Prospective cohort study/ NILS-LSA, elderly individuals, Japan | All-cause mortality | 1054 | 422 | n-6 intake<br>n-3 intake<br>EPA intake<br>DHA intake<br>Serum EPA<br>Serum DHA<br>Serum EPA/ARA | HR in tertiles (Highest to lowest; 95% CI) | 0.80 (0.59-1.07)<br>0.83 (0.61-1.12)<br>0.96 (0.71-1.30)<br>0.95 (0.70-1.28)<br>0.81 (0.60-1.09)<br>0.73 (0.53-0.99)<br>0.71 (0.53-0.96) | 0.13<br>0.22<br>0.78<br>0.72<br>0.17<br>0.047<br>0.02 |
| Zhuang et al.[4] | 2019 | Population based prospective cohort/ CHNS in China and NHANES in US | All-cause mortality | CHNS: 14,117<br>NHANES: 36,032 | 1,007<br>4,826 | Total PUFA intake<br><br>Omega-3 intake<br><br>Omega-6 intake<br><br>Omega 6/3 ratio | HR in quartiles (Highest to lowest; 95% CI) | China: 1.19 (0.93-1.52)<br>US: 0.86 (0.71-1.03)<br>China: 1.22 (1.00-1.50)<br>US: 0.85 (0.71-1.01)<br>China: 1.14 (0.89-1.47)<br>US: 0.84 (0.70-1.01)<br>China: 0.95 (0.80-1.14)<br>US: 0.99 (0.89-1.11) | 0.10<br>0.05<br>0.05<br>0.03<br>0.29<br>0.04<br>0.54<br>0.85 |
| *Circulating PUFAs* | | | | | | | | | |
| Wang et al.[5] | 2003 | An epidemiological survey of 65 rural countries in China | All-cause mortality | 6500 | NR | DHA<br>EPA<br>Total plasma Omega-3<br>Total plasma Omega-6 | Pearson correlation coefficient | Around -0.3 (mortality)<br>Around -0.2 (mortality)<br>-0.152<br>-0.001 | NR |

| | | | | | | | | | |
|---|---|---|---|---|---|---|---|---|---|
| Lindberg et al.[6] | 2008 | Prospective cohort/ Norway, elderly patients (mean=82.1), 3 y of follow-up | All-cause mortality | 254 | 101 | Plasma EPA | HR for quantile 1 (lowest, ref; 95% CI) compared with upper 3 quartiles combined | 0.52 (0.35-0.77) | NR |
| Chattipakorn et al.[7] | 2009 | Measure the heart tissues of cadavers with a history of CHD | - | 100 cadavers | - | Omega-3 (EPA + DHA)<br>Omega-6 (AA + LA) | P-value from two-sample Wilcoxon rank sum test (cardiac cause vs. noncardiac cause. Lower in the group with cardiac mortality) | Cadavers with heart disease<br>0.040<br>0.022 | NR |
| Lee et al.[8] | 2009 | Prospective cohort/ patients with acute myocardial infarction, 16.1 months follow-up | All-cause mortality | 508 | 36 | Plasma EPA | - | Lower plasma level of EPA was and independent predictor for all-cause mortality in female patients | NR |
| Pottala et al.[9] | 2010 | Prospective cohort/ outpatients with stable coronary heart disease, recruited between 2000-2002, San Francisco Bay Area | All-cause mortality | 956 | 237 | EPA + DHA in whole blood | HR above vs. below median (95% CI) | 0.74 (0.55-1.00) | 0.049 |
| Petrone et al.[10] | 2012 | Nested case-control/ US male physicians | Heart failure | 788 cases<br>788 matched controls | - | Plasma omega-6 PUFAs | OR in quartiles (Highest to lowest; 95% CI) | 0.87 (0.63-1.20) | 0.39 |
| Mozaffarian et al.[11] | 2013 | Prospective cohort/ older adults not taking fish oil supplements, 1992-2008, four US communities | All-cause mortality<br>CVD mortality | 2,692 | 1,625<br>570 | Plasma Total n-3 PUFA | HR in quintiles (Highest to lowest; 95% CI) | 0.73 (0.61-0.86)<br>0.65 (0.48-0.87) | <0.001<br>< 0.001 |
| Wu et al.[12] | 2014 | Community-based prospective cohort/ Cardiovascular Health Study, age>=65, US | All-cause mortality<br><br>CVD mortality | 2,792 | 1,994<br><br>678 | Plasma phospholipid n-6 PUFA<br>LA<br>(Others are NS) | HR in quintiles (Highest to lowest; 95% CI) | All-cause mortality<br>0.87 (0.74-1.02)<br>CVD mortality<br>0.78 (0.60-1.01) | <br>0.005<br><br>0.02 |
| Eide et al.[13] | 2016 | Cross-sectional study/ Renal transplant recipients, transplanted 1999-2011 | CVD mortality | 1990 | NR | plasma Marine n-3 PUFA levels | Per 1.0 wt% increase (95% CI) | 0.90 (0.82-0.98) | NR |
| Kleber et al.[14] | 2016 | Prospective cohort/ patients referred for coronary angiography, LURIC study, German | All-cause mortality<br><br>CVD mortality | 3259 | 975<br><br>614 | erythrocytes (red cells)<br>EPA<br><br>HS-Omega-3 Index | HR in tertiles (Highest to lowest; 95% CI) | EPA<br>All: 0.75 (0.64-0.88)<br>CVD: 0.70 (0.57-0.86)<br>HS-Omega-3 Index<br>All: 0.78 (0.67-0.92)<br>CVD: 0.78 (0.64-0.95) | <br>0.001<br>0.003<br><br>0.009<br>0.050 |
| Miura et al.[15] | 2016 | Population based prospective cohort/ Austria, drawn from Nambour Skin Cancer Study, in 1986 | All-cause mortality | 1,008 | 179<br>M: 98<br>F: 81 | Plasma total Omega-3<br><br><br>EPA<br><br><br>DPA<br><br><br>DHA<br><br><br>Total Omega-6 | HR per 1-SD increase (95% CI) | All: 0.96 (0.82-1.12)<br>M: 0.99 (0.80-1.22)<br>F: 0.95 (0.76-1.19)<br>All: 0.81 (0.69-0.95)<br>M: 0.78 (0.62-0.98)<br>F: 0.78 (0.65-0.94)<br>All: 0.90 (0.77-1.05)<br>M: 0.76 (0.60-0.97)<br>F: 0.98 (0.79-1.22)<br>All: 1.07 (0.92-1.25)<br>M: 1.12 (0.92-1.36)<br>F: 1.02 (0.82-1.28)<br>All: 0.87 (0.73-1.04)<br>M: 0.80 (0.92-1.04)<br>F: 0.96 (0.76-1.21) | 0.62<br>0.90<br>0.66<br>0.012<br>0.036<br>0.007<br>0.08<br>0.028<br>0.86<br>0.35<br>0.25<br>0.83<br>0.13<br>0.10<br>0.71 |
| Chen et al.[16] | 2016 | Meta-analysis/ general populations | All-cause mortality | 11 studies<br>371,965 | 31,185 | | Summary RR (Highest to lowest; 95% CI) | | NR |

| | | | | | | | | | |
|---|---|---|---|---|---|---|---|---|---|
| (Included Mozaffarian et al. and Bell et al. reports in the table) | | | | Dietary intakes (n=7) 361,273 Circulating levels (n=4) 10,692 | 27,624<br><br>3,561 | Circulating EPA<br>Circulating DHA | | 0.74 (0.60-0.90)<br>0.78 (0.64-0.93) | |
| Delgado et al.[17] | 2017 | Prospective cohort/ patients referred for coronary angiography, LURIC study, German | All-cause mortality<br>CVD mortality | 3259 | 975<br>614 | Plasma total omega-6 PUFA<br><br>GLA<br><br>ADA<br><br>DPA | 1 SD increase (95 % CI) | All: 0.93 (0.87-0.99)<br>CVD: 0.95 (0.87-1.03)<br><br>All: 0.88 (0.82-0.95)<br>CVD: 0.86 (0.79-0.95)<br>All: 1.10 (1.03-1.18)<br>CVD: 1.12 (1.04-1.22)<br>All: 1.12 (1.05-1.19)<br>CVD: 1.11 (1.02-1.20) | NR |
| Hamazaki et al.[18] | 2018 | Nested case-control study/ JPHC study, Japan | Coronary heart disease | 209 cases 418 matched controls | (168 myocardial infarction 41 sudden cardiac death) (157 non-fatal coronary events 52 fatal) | Plasma n-3 PUFAs | OR in quartiles (Highest to lowest; 95% CI) | CHD:0.79 (0.41-1.51)<br>MI: 0.91 (0.43-1.89)<br>Sudden death: 0.08 (0.01-0.88)<br>Non-fatal: 0.89 (0.42-1.89)<br>Fatal: 0.12 (0.02-0.75) | 0.51<br>0.90<br>0.04<br>0.97<br>0.03 |
| Harris et al.[19] | 2018 | Prospective cohort study/ the Framingham Heart Study Offspring cohort, US | All-cause mortality<br>CVD mortality<br>Cancer mortality<br>CVD events | 2500 | 350<br>58<br>146<br>245 | Omega-3 index red blood cells | HR in quintiles (Highest to lowest; 95% CI) | All-death: 0.66 (0.45-0.96)<br>CVD mortality: 0.39 (0.15-1.02)<br>Cancer mortality: 0.96 (0.56-1.64)<br>CVD events: 0.61 (0.37-0.99) | 0.02<br>0.10<br>0.88<br>0.008 |
| Kamleh et al.[20] | 2018 | Case-control study/ IMPROVE pan-European cohort, age 55-79 | Incidence of CVD events | Cases: 173 Matched controls: 172 Diabetics Cases: 40 Matched controls: 39 Non-diabetics Cases: 133 Matched controls: 133 | - | Plasma<br>DHA<br><br>GLA<br>AA<br>ADA | HR per 1-SD increase in log transformed metabolite concentration (95% CI) | Diabetics: NS<br>Non-diabetics:<br>DHA: 0.86 (0.72-1.02)<br><br>GLA: 0.72 (0.59-0.88)<br>AA: 0.83 (0.70-0.99)<br>ADA: 0.79 (0.66-0.95) | <br><br>0.085<br><br>0.001<br>0.037<br>0.012 |
| Miura et al.[21] | 2018 | Community-based prospective cohort/ women, 25-75 age, Australia | All-cause mortality | 564 | 81 | Plasma<br>Omega-3<br>Omega-6 | HR in tertiles (Highest to lowest; 95% CI) | Absolute (ug/ml)<br>Omega-3: 1.05 (0.57-1.92)<br>Omega-6: 1.19 (0.65-2.18)<br>Relative (%)<br>Omega-3: 0.97 (0.54-1.72)<br>Omega-6: 1.02 (0.58-1.78) | <br>0.84<br>0.60<br><br>0.77<br>1.00 |
| Marklund et al.[22] | 2019 | Meta-analysis | Total CVD<br><br>CVD mortality<br><br>Total CHD<br><br>Ischemic stroke | 30 prospective studies:<br><br>18 cohort<br><br>12 nested case-control or | 10,477<br><br>4,508<br><br>11,857<br><br>3,705 | Circulating<br>LA<br><br><br>AA | HR per interquintile range (95% CI) | LA<br>Total CVD: 0.93 (0.88-0.99)<br>CVD mortality: 0.78 (0.70-0.85)<br>Total CHD: 0.94 (0.88-1.00)<br>Ischemic stroke: 0.88 (0.79-0.98) | NR |

| | | | | | | | | | |
|---|---|---|---|---|---|---|---|---|---|
| | | | | case-control | | | | AA<br>Total CVD: 0.95 (0.90-1.01)<br>CVD mortality: 0.94 (0.86-1.02)<br>Total CHD: 0.99 (0.94-1.04)<br>Ischemic stroke: 0.99 (0.90-1.10) | |
| Harris et al.[23] | 2020 | Case-cohort study/ secondary analysis of ADVANCE study, patients with type 2 diabetes | Macrovascular disease<br>Microvascular disease<br>All-cause mortality<br>CVD mortality | 3,576 | 654<br>341<br>631<br>330 | Plasma<br>Omega-3<br>Omega-6 | HR per 1-SD increase of the percentage contribution of total fatty acids (95% CI) | Macrovascular disease<br>Omega-3: 0.87 (0.80-0.95)<br>Omega-6: 0.97 (0.89-1.07)<br>Microvascular disease<br>Omega-3: 1.01 (0.91-1.13)<br>Omega-6: 0.97 (0.86-1.10)<br>All-cause mortality<br>Omega-3: 0.91 (0.84-0.99)<br>Omega-6: 0.97 (0.88-1.07)<br>CVD mortality<br>Omega-3: 0.85 (0.75-0.96) | NR |
| Harris et al.[24] | 2021 | Meta-analysis/ age 50-81 | All-cause mortality<br>CVD mortality<br>Cancer mortality | All: 17 cohorts, 42,466<br>CVD: 15 cohorts<br>Cancer: 15 cohorts | 15,720<br><br><br>4,571<br><br>4,284 | Serum<br>EPA<br><br><br>DPA<br><br><br><br>DHA<br><br><br><br>EPA+DHA | HR in quintiles (Highest to lowest; 95% CI) | EPA<br>All: 0.82 (0.78-0.87)<br>CVD: 0.85 (0.77-0.94)<br>Cancer: 0.82 (0.74-0.91)<br>DPA<br>All: 0.84 (0.79-0.90)<br>CVD: 0.87 (0.78-0.98)<br>Cancer: 0.79 (0.70-0.90)<br>DHA<br>All: 0.85 (0.81-0.90)<br>CVD: 0.79 (0.72-0.88)<br>Cancer: 0.86 (0.78-0.95)<br>EPA+DHA<br>All: 0.84 (0.79-0.89)<br>CVD: 0.80 (0.73-0.88)<br>Cancer: 0.87 (0.78-0.96) | <br><0.001<br>0.006<br>0.008<br><br><0.001<br>0.16<br>0.008<br><br>0.01<br>0.002<br>0.06<br><br><0.001<br><0.001<br>0.06 |
| Kamalita et al.[25] | 2021 | Prospective cohort/ patients with prior MI, 60-80 years old, Dutch | All-cause mortality<br>CVD mortality | 4,067 | 1,877<br>834 | Circulating EPA + DHA | HR in quintiles (Highest to lowest; 95% CI) | 0.73 (0.63-0.86)<br>0.75 (0.60-0.95) | <0.001<br>0.016 |
| Diffenderfer et al.[26] | 2022 | Plasma fatty acid profiles in US | Heart disease mortality rate | 1,169,621 | NR | Plasma EPA<br>Omega-3 index | Pearson correlation coefficient | -0.504<br>-0.570 | <0.001<br><0.001 |
| *Dietary PUFAs* | | | | | | | | | |
| Bell et al.[27] | 2014 | Population based prospective cohort/ VITAL Study, 50-76 yrs old, western Washington State, recruited 2000-2002 | All-cause mortality<br>CVD mortality<br>Cancer mortality | 70,495 | 3,051<br>769<br>1,485 | EPA + DHA intake (diet + supplements) | HR in quartiles (Highest to lowest; 95% CI) | 0.82 (0.73-0.93)<br>0.87 (0.68-1.10)<br>0.77 (0.64-0.92) | 0.001<br>0.158<br>0.001 |
| Chen et al.[16] (Included Mozaffarian et al. and Bell et al. reports in the table) | 2016 | Meta-analysis/ general populations | All-cause mortality | 11 studies 371,965<br>Dietary intakes (n=7) 361,273<br>Circulating levels (n=4) 10,692 | 31,185<br><br>27,624<br><br>3,561 | N-3 LCPUFA intake | Summary RR (Highest to lowest; 95% CI) | 0.91 (0.84-0.98) | NR |

| | | | | | | | | | |
|---|---|---|---|---|---|---|---|---|---|
| Zhang et al.[28] | 2018 | Population based prospective cohort/ NIH-AARP Diet and Health Study, US | All-cause mortality<br><br><br>CVD mortality<br><br><br>Cancer mortality | 421,309<br>M: 240,729<br>F: 180,580 | All<br>M: 54,230<br>F: 30,882<br>CVD<br>M: 14,824<br>F: 7,541<br>Cancer<br>M: 20,041<br>F: 11,526 | Long-chain omega-3 PUFA intake | HR in quintiles (Highest to lowest; 95% CI) | All<br>M: 0.89 (0.86-0.92)<br>F: 0.90 (0.86-0.94)<br>CVD:<br>M: 0.85 (0.80-0.90)<br>F: 0.82 (0.75-0.90)<br>Cancer:<br>M: 0.95 (0.90-1.00)<br>F: 1.01 (0.93-1.09) | <br>< 0.001<br>< 0.001<br><br><0.001<br>< 0.001<br><br>0.04<br>0.51 |
| Kamalita et al.[25] | 2021 | Prospective cohort/ patients with prior MI, 60-80 years old, Dutch | All-cause mortality<br>CVD mortality | 4,067 | 1,877<br>834 | Dietary EPA + DHA intake | HR in quintiles (Highest to lowest; 95% CI) | All: 0.86 (0.75-0.99)<br>CVD: 0.84 (0.68-1.04) | 0.11<br>0.22 |
| *Fish intake and fish oil supplementation* | | | | | | | | | |
| Nagata et al.[29] | 2002 | Population based prospective cohort/ Japan, age >= 35, 1992-1999 | All-cause mortality<br><br><br>CVD mortality<br><br><br>Cancer mortality | 29,079<br>M: 13,355<br>F: 15,724 | 2,062<br>M: 1,163<br>F: 899<br>635<br>M: 308<br>F: 327<br>653<br>M: 400<br>F: 253 | Fish oil | HR in quintiles (Highest to lowest; 95% CI) | M: 0.87 (0.73-1.05)<br>F: 0.77 (0.62-0.94)<br><br>M: 0.76 (0.54-1.07)<br>F: 0.77 (0.55-1.00)<br><br>M: 0.89 (0.66-1.20)<br>F: 0.70 (0.47-1.05) | 0.38<br>0.01<br><br>0.27<br>0.16<br><br>0.52<br>0.15 |
| Li et al.[30] | 2020 | Population based prospective cohort/ UK biobank | All-cause mortality<br>CVD mortality | 427,678 | 12,928<br>3282 | Fish oil supplementation | HR (Yes to No; 95% CI) | 0.87 (0.83-0.90)<br>0.84 (0.78-0.91) | NR |
| Jayedi et al.[31] | 2021 | Meta-analysis/ patients with type 2 diabetes | <br><br>All-cause mortality<br>CVD diseases<br>CHD<br>MI<br>stroke | Total:<br>9 studies<br>(57,394)<br>8 studies<br>(57,077)<br>4 studies<br>(8781) | NR<br><br><br><br>791<br>376<br>372 | Fish consumption | Pooled RR (Highest to lowest; 95% CI) | <br><br>0.86 (0.76-0.96)<br><br>0.61 (0.29-0.93)<br>NS<br>NS | NR |
| Liu et al.[32] | 2022 | Population based prospective cohort/ UK Biobank, UK | Overall cancer | 470,804 | 28,417 | Fish oil supplementation | HR (Yes to No; 95% CI) | 0.97 (0.95-1.00) | 0.06 |
| Ma et al.[33] | 2022 | Prospective cohort/ patients with hypertension, UK Biobank, UK | Incidence of cardiometabolic multimorbidity (CMM)<br>All-cause mortality<br>CVD mortality<br>Cancer mortality | 81,579 | 15,990<br><br>6,456<br>1,308<br>3,307 | Fish oil supplementation | HR (Yes to No; 95% CI) | 0.92 (0.89-0.96)<br><br><br>0.90 (0.85-0.95)<br>0.86 (0.76-0.98)<br>0.99 (0.91-1.07) | <0.001<br><br><br><0.001<br>0.027<br>0.742 |
| Abbreviations: HR, hazards ratio; NR, not recorded; NS, not significant. | | | | | | | | | |
