## Supplementary Figures and Tables for "Higher ratio of plasma omega-6/omega-3 fatty acids is associated with greater risk of all-cause, cancer, and cardiovascular mortality: a population-based cohort study in UK Biobank"

**Table S2.** Risk estimates^a^ of ratio of plasma omega-6 to omega-3 PUFAs with all-cause, cancer and CVD mortality, stratified by potential risk factors, in the UK Biobank Study (n = 85,425).

| **Stratification variables and ratio quintiles** | **Causes of death** | | | | | | | |
| --- | --- | --- | --- | --- | --- | --- | --- | --- |
|  | **All-cause** | |  | **Cancer** | |  | **Cardiovascular diseases** | |
|  | Death (n) | HR (95% CI) |  | Death (n) | HR (95% CI) |  | Death (n) | HR (95% CI) |
| **Age, years** |  |  |  |  |  |  |  |  |
| **Continuous (p for interaction)** | 0.224 | |  | 0.118 | |  | 0.875 | |
| **< 58** |  |  |  |  |  |  |  |  |
| 1 | 194 | 1.00 (ref) |  | 85 | 1.00 (ref) |  | 56 | 1.00 (ref) |
| 2 | 223 | 0.92 (0.74-1.14) |  | 118 | 1.08 (0.79-1.47) |  | 47 | 0.74 (0.47-1.14) |
| 3 | 284 | 0.92 (0.75-1.13) |  | 129 | 0.92 (0.68-1.26) |  | 79 | 0.98 (0.66-1.45) |
| 4 | 305 | 0.93 (0.76-1.14) |  | 154 | 1.05 (0.78-1.41) |  | 71 | 0.80 (0.53-1.20) |
| 5 | 430 | 1.11 (0.91-1.34) |  | 188 | 1.07 (0.80-1.43) |  | 112 | 1.11 (0.77-1.61) |
| *P for trend* |  | *0.070* |  |  | *0.611* |  |  | *0.193* |
| **≥ 58** |  |  |  |  |  |  |  |  |
| 1 | 1154 | 1.00 (ref) |  | 508 | 1.00 (ref) |  | 313 | 1.00 (ref) |
| 2 | 1033 | 0.95 (0.86-1.04) |  | 445 | 0.94 (0.81-1.08) |  | 268 | 0.92 (0.76-1.11) |
| 3 | 952 | 0.98 (0.89-1.09) |  | 414 | 0.96 (0.83-1.11) |  | 242 | 0.92 (0.75-1.12) |
| 4 | 947 | 1.06 (0.96-1.17) |  | 394 | 1.06 (0.91-1.23) |  | 235 | 1.03 (0.84-1.25) |
| 5 | 939 | 1.17 (1.06-1.29) |  | 359 | 1.04 (0.89-1.21) |  | 245 | 1.24 (1.02-1.51) |
| *P for trend* |  | *< 0.001* |  |  | *0.317* |  |  | *0.010* |
| *P for interaction* |  | *0.798* |  |  | *0.853* |  |  | *0.588* |
| **Sex** |  |  |  |  |  |  |  |  |
| **Male** |  |  |  |  |  |  |  |  |
| 1 | 730 | 1.00 (ref) |  | 287 | 1.00 (ref) |  | 236 | 1.00 (ref) |
| 2 | 755 | 0.99 (0.89-1.11) |  | 317 | 1.08 (0.91-1.29) |  | 217 | 0.88 (0.72-1.09) |
| 3 | 777 | 1.01 (0.90-1.13) |  | 301 | 0.99 (0.83-1.19) |  | 227 | 0.91 (0.73-1.12) |
| 4 | 834 | 1.11 (0.99-1.24) |  | 338 | 1.15 (0.96-1.37) |  | 224 | 1.02 (0.83-1.25) |
| 5 | 980 | 1.29 (1.16-1.44) |  | 355 | 1.17 (0.98-1.40) |  | 282 | 1.31 (1.08-1.60) |
| *P for trend* |  | *< 0.001* |  |  | *0.057* |  |  | *< 0.001* |
| **Female** |  |  |  |  |  |  |  |  |
| 1 | 618 | 1.00 (ref) |  | 306 | 1.00 (ref) |  | 133 | 1.00 (ref) |
| 2 | 501 | 0.91 (0.79-1.05) |  | 246 | 0.85 (0.70-1.04) |  | 98 | 0.92 (0.66-1.27) |
| 3 | 459 | 1.03 (0.89-1.19) |  | 242 | 1.00 (0.81-1.22) |  | 94 | 1.18 (0.85-1.64) |
| 4 | 418 | 1.07 (0.92-1.25) |  | 210 | 1.08 (0.88-1.33) |  | 82 | 1.07 (0.76-1.52) |
| 5 | 389 | 1.22 (1.04-1.42) |  | 192 | 1.11 (0.89-1.38) |  | 75 | 1.31 (0.91-1.88) |
| *P for trend* |  | *0.003* |  |  | *0.110* |  |  | *0.099* |
| *P for interaction* |  | *0.798* |  |  | *0.333* |  |  | *0.731* |
| **TDI** |  |  |  |  |  |  |  |  |
| **Continuous (p for interaction)** | 0.196 | |  | 0.351 | |  | 0.945 | |
| **< -2** |  |  |  |  |  |  |  |  |
| 1 | 719 | 1.00 (ref) |  | 352 | 1.00 (ref) |  | 180 | 1.00 (ref) |
| 2 | 582 | 0.94 (0.83-1.06) |  | 282 | 0.88 (0.74-1.05) |  | 129 | 0.85 (0.66-1.11) |
| 3 | 574 | 1.00 (0.88-1.13) |  | 281 | 0.92 (0.77-1.10) |  | 139 | 1.03 (0.80-1.34) |
| 4 | 549 | 1.07 (0.95-1.22) |  | 268 | 1.04 (0.87-1.24) |  | 128 | 1.11 (0.85-1.44) |
| 5 | 500 | 1.17 (1.02-1.33) |  | 240 | 1.06 (0.88-1.28) |  | 121 | 1.35 (1.04-1.75) |
| *P for trend* |  | *0.004* |  |  | *0.240* |  |  | *0.005* |
| **≥ -2** |  |  |  |  |  |  |  |  |
| 1 | 629 | 1.00 (ref) |  | 241 | 1.00 (ref) |  | 189 | 1.00 (ref) |
| 2 | 673 | 0.99 (0.87-1.12) |  | 280 | 1.14 (0.93-1.39) |  | 186 | 0.93 (0.73-1.18) |
| 3 | 662 | 1.05 (0.93-1.20) |  | 262 | 1.11 (0.91-1.36) |  | 182 | 0.95 (0.74-1.21) |
| 4 | 702 | 1.13 (1.00-1.28) |  | 280 | 1.25 (1.02-1.53) |  | 178 | 0.98 (0.77-1.25) |
| 5 | 868 | 1.38 (1.22-1.55) |  | 307 | 1.29 (1.06-1.58) |  | 235 | 1.32 (1.05-1.66) |
| *P for trend* |  | *< 0.001* |  |  | 0.010 |  |  | *0.004* |
| *P for interaction* |  | *0.268* |  |  | *0.395* |  |  | *0.846* |
| **BMI** | 0.925 | |  | 0.822 | |  | 0.298 | |
| **Continuous (p for interaction)** |  |  |  |  |  |  |  |  |
| **< 25** |  |  |  |  |  |  |  |  |
| 1 | 373 | 1.00 (ref) |  | 179 | 1.00 (ref) |  | 79 | 1.00 (ref) |
| 2 | 298 | 0.90 (0.76-1.07) |  | 139 | 0.91 (0.71-1.16) |  | 66 | 0.92 (0.63-1.33) |
| 3 | 305 | 0.93 (0.78-1.11) |  | 144 | 0.85 (0.66-1.10) |  | 59 | 0.89 (0.61-1.31) |
| 4 | 333 | 0.98 (0.83-1.17) |  | 151 | 1.05 (0.82-1.34) |  | 66 | 0.87 (0.59-1.27) |
| 5 | 429 | 1.15 (0.97-1.36) |  | 184 | 1.04 (0.81-1.34) |  | 97 | 1.32 (0.93-1.88) |
| *P for trend* |  | *0.025* |  |  | *0.399* |  |  | *0.071* |
| **≥ 25** |  |  |  |  |  |  |  |  |
| 1 | 965 | 1.00 (ref) |  | 413 | 1.00 (ref) |  | 286 | 1.00 (ref) |
| 2 | 948 | 0.99 (0.89-1.10) |  | 422 | 1.02 (0.87-1.18) |  | 244 | 0.90 (0.73-1.10) |
| 3 | 917 | 1.05 (0.95-1.17) |  | 396 | 1.06 (0.91-1.24) |  | 261 | 1.02 (0.83-1.24) |
| 4 | 907 | 1.14 (1.03-1.26) |  | 392 | 1.15 (0.98-1.35) |  | 236 | 1.09 (0.89-1.33) |
| 5 | 927 | 1.30 (1.17-1.44) |  | 362 | 1.19 (1.01-1.40) |  | 257 | 1.30 (1.07-1.59) |
| *P for trend* |  | *< 0.001* |  |  | 0.012 |  |  | 0.001 |
| *P for interaction* |  | *0.598* |  |  | *0.782* |  |  | *0.615* |
| **Comorbidities** |  |  |  |  |  |  |  |  |
| **Yes** |  |  |  |  |  |  |  |  |
| 1 | 878 | 1.00 (ref) |  | 329 | 1.00 (ref) |  | 285 | 1.00 (ref) |
| 2 | 806 | 0.95 (0.85-1.06) |  | 302 | 0.99 (0.83-1.18) |  | 239 | 0.86 (0.70-1.05) |
| 3 | 740 | 0.95 (0.85-1.06) |  | 270 | 0.95 (0.79-1.14) |  | 220 | 0.84 (0.67-1.03) |
| 4 | 797 | 1.09 (0.98-1.22) |  | 276 | 1.05 (0.87-1.26) |  | 242 | 1.09 (0.89-1.33) |
| 5 | 849 | 1.27 (1.14-1.42) |  | 282 | 1.12 (0.93-1.35) |  | 250 | 1.24 (1.01-1.52) |
| *P for trend* |  | *< 0.001* |  |  | 0.172 |  |  | 0.002 |
| **No** |  |  |  |  |  |  |  |  |
| 1 | 470 | 1.00 (ref) |  | 264 | 1.00 (ref) |  | 84 | 1.00 (ref) |
| 2 | 450 | 0.97 (0.84-1.12) |  | 261 | 0.98 (0.80-1.18) |  | 76 | 0.99 (0.70-1.42) |
| 3 | 496 | 1.12 (0.97-1.29) |  | 273 | 1.05 (0.86-1.27) |  | 101 | 1.37 (0.98-1.91) |
| 4 | 455 | 1.09 (0.94-1.26) |  | 272 | 1.20 (0.99-1.45) |  | 64 | 0.82 (0.56-1.20) |
| 5 | 520 | 1.24 (1.07-1.43) |  | 265 | 1.17 (0.96-1.43) |  | 107 | 1.47 (1.05-2.06) |
| *P for trend* |  | *0.001* |  |  | *0.027* |  |  | *0.046* |
| *P for interaction* |  | *0.219* |  |  | *0.736* |  |  | *0.006* |
| **Physical activity** |  |  |  |  |  |  |  |  |
| **Low or moderate** |  |  |  |  |  |  |  |  |
| 1 | 686 | 1.00 (ref) |  | 303 | 1.00 (ref) |  | 175 | 1.00 (ref) |
| 2 | 626 | 0.96 (0.86-1.07) |  | 271 | 0.97 (0.82-1.14) |  | 158 | 0.93 (0.74-1.15) |
| 3 | 582 | 1.01 (0.90-1.13) |  | 251 | 1.01 (0.85-1.19) |  | 136 | 0.89 (0.71-1.12) |
| 4 | 617 | 1.11 (1.00-1.24) |  | 267 | 1.12 (0.95-1.33) |  | 162 | 1.10 (0.88-1.36) |
| 5 | 651 | 1.30 (1.17-1.46) |  | 247 | 1.17 (0.98-1.39) |  | 180 | 1.37 (1.10-1.70) |
| *P for trend* |  | *< 0.001* |  |  | 0.020 |  |  | *< 0.001* |
| **High** |  |  |  |  |  |  |  |  |
| 1 | 388 | 1.00 (ref) |  | 183 | 1.00 (ref) |  | 100 | 1.00 (ref) |
| 2 | 346 | 0.95 (0.82-1.10) |  | 172 | 1.01 (0.82-1.24) |  | 77 | 0.82 (0.61-1.11) |
| 3 | 352 | 1.04 (0.90-1.21) |  | 157 | 0.98 (0.79-1.22) |  | 99 | 1.15 (0.87-1.53) |
| 4 | 341 | 1.06 (0.91-1.23) |  | 168 | 1.10 (0.89-1.36) |  | 76 | 0.92 (0.68-1.25) |
| 5 | 383 | 1.20 (1.04-1.39) |  | 165 | 1.11 (0.89-1.38) |  | 100 | 1.22 (0.91-1.62) |
| *P for trend* |  | 0.003 |  |  | *0.228* |  |  | *0.099* |
| *P for interaction* |  | *0.883* |  |  | *0.994* |  |  | *0.188* |
| **Smoke status** |  |  |  |  |  |  |  |  |
| **Yes** |  |  |  |  |  |  |  |  |
| 1 | 155 | 1.00 (ref) |  | 65 | 1.00 (ref) |  | 41 | 1.00 (ref) |
| 2 | 193 | 1.01 (0.79-1.30) |  | 82 | 0.95 (0.65-1.37) |  | 56 | 1.47 (0.90-2.38) |
| 3 | 249 | 1.22 (0.96-1.54) |  | 96 | 0.98 (0.68-1.42) |  | 71 | 1.53 (0.95-2.47) |
| 4 | 292 | 1.24 (0.98-1.56) |  | 132 | 1.23 (0.87-1.74) |  | 74 | 1.55 (0.97-2.49) |
| 5 | 438 | 1.57 (1.26-1.95) |  | 176 | 1.41 (1.01-1.97) |  | 116 | 2.09 (1.33-3.27) |
| *P for trend* |  | *< 0.001* |  |  | *0.003* |  |  | *0.001* |
| **No** |  |  |  |  |  |  |  |  |
| 1 | 1178 | 1.00 (ref) |  | 523 | 1.00 (ref) |  | 322 | 1.00 (ref) |
| 2 | 1054 | 0.96 (0.87-1.05) |  | 479 | 0.99 (0.86-1.14) |  | 256 | 0.83 (0.69-1.00) |
| 3 | 973 | 0.98 (0.89-1.08) |  | 441 | 1.00 (0.86-1.15) |  | 247 | 0.90 (0.74-1.09) |
| 4 | 949 | 1.06 (0.97-1.17) |  | 413 | 1.09 (0.94-1.26) |  | 231 | 0.96 (0.79-1.17) |
| 5 | 918 | 1.18 (1.06-1.30) |  | 367 | 1.06 (0.91-1.23) |  | 238 | 1.18 (0.97-1.43) |
| *P for trend* |  | *< 0.001* |  |  | *0.264* |  |  | *0.024* |
| *P for interaction* |  | *0.007* |  |  | *0.125* |  |  | *0.116* |

Abbreviations: CI, confidence interval; HR, hazards ratio; ref, reference.

^a^ From Cox proportional hazards regression; adjusted for age (years; continuous), sex (male, female), race (White, Black, Asian, Others), Townsend deprivation index (continuous), assessment centre, BMI (kg/m2; continuous), smoking status (never, previous, current), alcohol intake status (never, previous, current), physical activity (low, moderate, high), and comorbidities (yes, no).

**Table S3.** Associations^a^ of plasma omega-3 PUFAs percentage with all-cause, cancer, and CVD mortality risk in the UK Biobank Study.

| **Omega ratio variable forms** | **Causes of death** | | | | | | | | | | | | | |
| --- | --- | --- | --- | --- | --- | --- | --- | --- | --- | --- | --- | --- | --- | --- |
|  | **All-cause** | | | |  | **Cancer** | | | |  | **Cardiovascular diseases** | | | |
|  | Death # | Model 1^b^ | Model 2^c^ | Model 3^d^ |  | Death # | Model 1^b^ | Model 2^c^ | Model 3^d^ |  | Death # | Model 1^b^ | Model 2^c^ | Model 3^d^ |
|  |  | HR  (95% CI) | HR  (95% CI) | HR  (95% CI) |  |  | HR  (95% CI) | HR  (95% CI) | HR  (95% CI) |  |  | HR  (95% CI) | HR  (95% CI) | HR  (95% CI) |
| Continuous | 6,461 | 0.91 | 0.90 | 0.94 |  | 2,794 | 0.93 | 0.92 | 0.95 |  | 1,668 | 0.92 | 0.90 | 0.93 |
|  |  | (0.89-0.92) | (0.88-0.91) | (0.92-0.95) |  |  | (0.90-0.95) | (0.89-0.94) | (0.93-0.98) |  |  | (0.89-0.95) | (0.87-0.94) | (0.90-0.97) |
| Quintiles (median) | |  |  |  |  |  |  |  |  |  |  |  |  |  |
| 1 (2.7) | 1,526 | 1.00 | 1.00 | 1.00 |  | 600 | 1.00 | 1.00 | 1.00 |  | 413 | 1.00 | 1.00 | 1.00 |
|  |  | (ref) | (ref) | (ref) |  |  | (ref) | (ref) | (ref) |  |  | (ref) | (ref) | (ref) |
| 2 (3.5) | 1,283 | 0.81 | 0.78 | 0.82 |  | 542 | 0.85 | 0.83 | 0.89 |  | 327 | 0.77 | 0.74 | 0.75 |
|  |  | (0.75-0.87) | (0.72-0.84) | (0.76-0.90) |  |  | (0.76-0.96) | (0.74-0.94) | (0.78-1.01) |  |  | (0.67-0.90) | (0.64-0.86) | (0.64-0.89) |
| 3 (4.2) | 1,246 | 0.75 | 0.72 | 0.77 |  | 581 | 0.86 | 0.84 | 0.91 |  | 300 | 0.69 | 0.65 | 0.67 |
|  |  | (0.70-0.81) | (0.67-0.78) | (0.71-0.84) |  |  | (0.77-0.97) | (0.75-0.94) | (0.79-1.04) |  |  | (0.59-0.80) | (0.56-0.75) | (0.56-0.80) |
| 4 (4.9) | 1,244 | 0.71 | 0.68 | 0.76 |  | 553 | 0.77 | 0.75 | 0.84 |  | 323 | 0.73 | 0.68 | 0.72 |
|  |  | (0.66-0.77) | (0.63-0.73) | (0.69-0.83) |  |  | (0.69-0.87) | (0.66-0.84) | (0.73-0.96) |  |  | (0.63-0.85) | (0.59-0.79) | (0.60-0.85) |
| 5 (6.3) | 1,162 | 0.62 | 0.59 | 0.69 |  | 518 | 0.67 | 0.64 | 0.75 |  | 305 | 0.65 | 0.61 | 0.68 |
|  |  | (0.57-0.67) | (0.54-0.64) | (0.63-0.76) |  |  | (0.59-0.76) | (0.57-0.73) | (0.65-0.87) |  |  | (0.56-0.76) | (0.52-0.71) | (0.57-0.82) |
| *P*_trend_ |  | *<0.001* | *<0.001* | *<0.001* |  |  | *<0.001* | *<0.001* | *<0.001* |  |  | *<0.001* | *<0.001* | *<0.001* |

Abbreviations: CI, confidence interval; HR, hazards ratio; ref, reference.

^a^ From Cox proportional hazards regression.

^b^ Adjusted for age (years; continuous), sex (male, female), race (White, Black, Asian, Others), Townsend deprivation index (continuous), assessment centre.

^c^ Adjusted for omega-6, age (years; continuous), sex (male, female), race (White, Black, Asian, Others), Townsend deprivation index (continuous), assessment centre.

^d^ Adjusted for omega-6, age (years; continuous), sex (male, female), race (White, Black, Asian, Others), Townsend deprivation index (continuous), assessment centre, BMI (kg/m2; continuous), smoking status (never, previous, current), alcohol intake status (never, previous, current), physical activity (low, moderate, high), and comorbidities (yes, no).

**Table S4.** Associations^a^ of plasma omega-6 PUFAs percentage with all-cause, cancer, and CVD mortality risk in the UK Biobank Study.

| **Omega ratio variable forms** | **Causes of death** | | | | | | | | | | | | | |
| --- | --- | --- | --- | --- | --- | --- | --- | --- | --- | --- | --- | --- | --- | --- |
|  | **All-cause** | | | |  | **Cancer** | | | |  | **Cardiovascular diseases** | | | |
|  | Death # | Model 1^b^ | Model 2^c^ | Model 3^d^ |  | Death # | Model 1^b^ | Model 2^c^ | Model 3^d^ |  | Death # | Model 1^b^ | Model 2^c^ | Model 3^d^ |
|  |  | HR  (95% CI) | HR  (95% CI) | HR  (95% CI) |  |  | HR  (95% CI) | HR  (95% CI) | HR  (95% CI) |  |  | HR  (95% CI) | HR  (95% CI) | HR  (95% CI) |
| Continuous | 6,461 | 0.96 | 0.96 | 0.98 |  | 2,794 | 0.97 | 0.97 | 0.98 |  | 1,668 | 0.94 | 0.94 | 0.99 |
|  |  | (0.95-0.97) | (0.95-0.96) | (0.97-0.99) |  |  | (0.96-0.98) | (0.96-0.98) | (0.97-1.00) |  |  | (0.93-0.96) | (0.93-0.95) | (0.97-1.00) |
| Quintiles (median) | |  |  |  |  |  |  |  |  |  |  |  |  |  |
| 1 (33) | 1,911 | 1.00 | 1.00 | 1.00 |  | 772 | 1.00 | 1.00 | 1.00 |  | 545 | 1.00 | 1.00 | 1.00 |
|  |  | (ref) | (ref) | (ref) |  |  | (ref) | (ref) | (ref) |  |  | (ref) | (ref) | (ref) |
| 2 (37) | 1,426 | 0.82 | 0.82 | 0.90 |  | 628 | 0.86 | 0.86 | 0.91 |  | 363 | 0.76 | 0.76 | 0.87 |
|  |  | (0.76-0.88) | (0.77-0.88) | (0.83-0.97) |  |  | (0.77-0.96) | (0.78-0.96) | (0.80-1.02) |  |  | (0.66-0.87) | (0.67-0.87) | (0.74-1.02) |
| 3 (39) | 1,259 | 0.78 | 0.78 | 0.95 |  | 532 | 0.78 | 0.78 | 0.87 |  | 329 | 0.76 | 0.75 | 1.09 |
|  |  | (0.73-0.84) | (0.73-0.84) | (0.87-1.03) |  |  | (0.70-0.87) | (0.70-0.87) | (0.77-0.99) |  |  | (0.66-0.87) | (0.66-0.87) | (0.92-1.28) |
| 4 (40) | 1,075 | 0.75 | 0.73 | 0.90 |  | 502 | 0.82 | 0.80 | 0.93 |  | 249 | 0.65 | 0.63 | 0.98 |
|  |  | (0.69-0.81) | (0.68-0.79) | (0.83-0.99) |  |  | (0.73-0.92) | (0.71-0.90) | (0.81-1.06) |  |  | (0.56-0.76) | (0.55-0.74) | (0.82-1.17) |
| 5 (42) | 790 | 0.64 | 0.60 | 0.77 |  | 360 | 0.69 | 0.65 | 0.80 |  | 182 | 0.53 | 0.50 | 0.83 |
|  |  | (0.59-0.69) | (0.55-0.65) | (0.70-0.85) |  |  | (0.61-0.78) | (0.57-0.74) | (0.68-0.92) |  |  | (0.45-0.63) | (0.42-0.59) | (0.68-1.02) |
| *P*_trend_ |  | *<0.001* | *<0.001* | *<0.001* |  |  | *<0.001* | *<0.001* | *0.007* |  |  | *<0.001* | *<0.001* | *0.363* |

Abbreviations: CI, confidence interval; HR, hazards ratio; ref, reference.

^a^ From Cox proportional hazards regression.

^b^ Adjusted for age (years; continuous), sex (male, female), race (White, Black, Asian, Others), Townsend deprivation index (continuous), assessment centre.

^c^ Adjusted for omega-3, age (years; continuous), sex (male, female), race (White, Black, Asian, Others), Townsend deprivation index (continuous), assessment centre.

^d^ Adjusted for omega-3, age (years; continuous), sex (male, female), race (White, Black, Asian, Others), Townsend deprivation index (continuous), assessment centre, BMI (kg/m2; continuous), smoking status (never, previous, current), alcohol intake status (never, previous, current), physical activity (low, moderate, high), and comorbidities (yes, no).

**Table S5.** Fully adjusted joint associations^a^ of plasma omega-3 PUFAs percentage and omega-6 PUFAs percentage with all-cause and cause-specific mortality in the UK Biobank Study.

|  | Omega-6% quintiles | | | | | | | | | |
| --- | --- | --- | --- | --- | --- | --- | --- | --- | --- | --- |
|  | All-cause mortality^b^ | | | | | | | | | |
| Omega-3% quintiles | Death/Total | 1  HR (95%CI) | Death/Total | 2  HR (95%CI) | Death/Total | 3  HR (95%CI) | Death/Total | 4  HR (95%CI) | Death/Total | 5  HR (95%CI) |
| 1 | 391/2,830 | 1.00 (Ref)^c^ | 304/2,693 | 0.96 (0.80-1.15) | 289/2,955 | 0.97 (0.81-1.17) | 282/3,444 | 0.93 (0.76-1.12) | 260/5,163 | 0.73 (0.59-0.89) |
| 2 | 370/3,483 | 0.75 (0.64-0.89) | 279/3,092 | 0.80 (0.66-0.97) | 253/3,144 | 0.87 (0.72-1.06) | 220/3,420 | 0.76 (0.61-0.94) | 161/3,946 | 0.64 (0.50-0.81) |
| 3 | 401/3,600 | 0.81 (0.69-0.96) | 265/3,390 | 0.66 (0.54-0.80) | 236/3,362 | 0.70 (0.56-0.86) | 193/3,463 | 0.62 (0.49-0.78) | 151/3,270 | 0.66 (0.82-0.85) |
| 4 | 396/3,598 | 0.79 (0.67-0.94) | 292/3,680 | 0.70 (0.58-0.85) | 222/3,622 | 0.60 (0.48-0.75) | 195/3,428 | 0.69 (0.54-0.87) | 139/2,757 | 0.73 (0.56-0.95) |
| 5 | 353/3,574 | 0.68 (0.57-0.82) | 286/4,230 | 0.55 (0.45-0.68) | 259/4,002 | 0.68 (0.54-0.85) | 185/3,330 | 0.60 (0.47-0.77) | 79/1,949 | 0.48 (0.35-0.67)^d^ |
|  | Cancer mortality^b^ | | | | | | | | | |
| 1 | 139/2,830 | 1.00 (Ref)^c^ | 122/2,693 | 1.02 (0.77-1.36) | 98/2,955 | 0.76 (0.55-1.03) | 131/3,444 | 1.03 (0.76-1.40) | 110/5,163 | 0.72 (0.52-1.00) |
| 2 | 132/3,483 | 0.72 (0.55-0.96) | 117/3,092 | 0.87 (0.65-1.17) | 114/3,144 | 0.96 (0.72-1.30) | 106/3,420 | 0.81 (0.59-1.13) | 73/3,946 | 0.64 (0.44-0.93) |
| 3 | 182/3,600 | 0.98 (0.76-1.27) | 118/3,390 | 0.73 (0.54-0.99) | 113/3,362 | 0.74 (0.53-1.03) | 92/3,463 | 0.70 (0.49-0.99) | 76/3,270 | 0.72 (0.49-1.05) |
| 4 | 168/3,598 | 0.93 (0.71-1.21) | 136/3,680 | 0.73 (0.54-1.00) | 95/3,622 | 0.60 (0.43-0.84) | 91/3,428 | 0.68 (0.47-0.99) | 63/2,757 | 0.74 (0.49-1.11) |
| 5 | 151/3,574 | 0.74 (0.56-0.99) | 135/4,230 | 0.63 (0.46-0.87) | 112/4,002 | 0.63 (0.44-0.91) | 82/3,330 | 0.58 (0.39-0.86) | 38/1,949 | 0.53 (0.33-0.86)^d^ |
|  | CVD mortality^b^ | | | | | | | | | |
| 1 | 116/2,830 | 1.00 (Ref)^c^ | 82/2,693 | 0.97 (0.69-1.36) | 81/2,955 | 1.15 (0.82-1.61) | 66/3,444 | 0.95 (0.65-1.37) | 68/5,163 | 0.86 (0.59-1.26) |
| 2 | 114/3,483 | 0.73 (0.54-1.00) | 71/3,092 | 0.77 (0.53-1.10) | 61/3,144 | 0.88 (0.61-1.27) | 45/3,420 | 0.79 (0.51-1.22) | 36/3,946 | 0.68 (0.43-1.09) |
| 3 | 100/3,600 | 0.70 (0.51-0.97) | 64/3,390 | 0.52 (0.35-0.78) | 63/3,362 | 0.73 (0.50-1.07) | 46/3,463 | 0.72 (0.46-1.13) | 27/3,270 | 0.56 (0.33-0.95) |
| 4 | 119/3,598 | 0.72 (0.52-0.98) | 66/3,680 | 0.65 (0.44-0.96) | 61/3,622 | 0.66 (0.43-1.01) | 50/3,428 | 0.80 (0.51-1.27) | 27/2,757 | 0.73 (0.42-1.24) |
| 5 | 96/3,574 | 0.63 (0.45-0.89) | 80/4,230 | 0.56 (0.37-0.85) | 63/4,002 | 0.76 (0.49-1.17) | 42/3,330 | 0.75 (0.46-1.21) | 24/1,949 | 0.71 (0.38-1.31)^d^ |

Abbreviations: CI, confidence interval; HR, hazards ratio; ref, reference; omega-3%, omega-3 fatty acids to total fatty acids percentage; omega-6%, omega-6 fatty acids to total fatty acids percentage.

^a^ From Cox proportional hazards regression.

^b^ Adjusted for age (years; continuous), sex (male, female), race (White, Black, Asian, Others), Townsend deprivation index (continuous), assessment centre, BMI (kg/m2; continuous), smoking status (never, previous, current), alcohol intake status (never, previous, current), physical activity (low, moderate, high), and comorbidities (yes, no).

^c^ Reference category: participants who had both low omega-3 fatty acids percentage and omega-6 fatty acids percentage.

^d^ *P*_interaction_ for all-cause, cancer, and CVD mortality 0.04, 0.15, and 0.98, respectively.

**Table S6.** Associations of the dietary PUFAs with all-cause, cancer, and CVD mortality risk in the UK Biobank (n=153,064).

|  | **Dietary Omega-3%^d^** | | |  | **Dietary Omega-6%^e^** | | |  | **Dietary Omega-ratio^f^** | | |
| --- | --- | --- | --- | --- | --- | --- | --- | --- | --- | --- | --- |
| **Cause of death** | All-cause | Cancer | CVD |  | All-cause | Cancer | CVD |  | All-cause | Cancer | CVD |
|  | HR  (95% CI) | HR  (95% CI) | HR  (95% CI) |  | HR  (95% CI) | HR  (95% CI) | HR  (95% CI) |  | HR  (95% CI) | HR  (95% CI) | HR  (95% CI) |
| Continuous | 0.98  (0.96-1.00) | 0.96  (0.93-0.98) | 1.00  (0.96-1.04) |  | 1.00  (0.99-1.00) | 1.00  (0.99-1.01) | 1.00  (0.99-1.01) |  | 1.01  (1.00-1.02) | 1.03  (1.01-1.05) | 0.99  (0.96-1.02) |
| Quintiles^a,b,c^ |  |  |  |  |  |  |  |  |  |  |  |
| 1 | 1.00 (ref) | 1.00 (ref) | 1.00 (ref) |  | 1.00 (ref) | 1.00 (ref) | 1.00 (ref) |  | 1.00 (ref) | 1.00 (ref) | 1.00 (ref) |
| 2 | 0.92  (0.85-0.99) | 0.93  (0.84-1.03) | 0.99  (0.84-1.16) |  | 0.87  (0.81-0.94) | 0.92  (0.83-1.02) | 0.87  (0.74-1.02) |  | 0.99  (0.92-1.07) | 0.99  (0.88-1.10) | 0.96  (0.82-1.13) |
| 3 | 0.96  (0.89-1.04) | 0.95  (0.85-1.06) | 1.09  (0.92-1.29) |  | 0.91  (0.84-0.98) | 0.97  (0.87-1.07) | 0.92  (0.78-1.07) |  | 1.00  (0.92-1.08) | 1.01  (0.90-1.12) | 0.93  (0.79-1.09) |
| 4 | 0.90  (0.82-0.97) | 0.85  (0.75-0.95) | 1.02  (0.85-1.21) |  | 0.90  (0.84-0.98) | 0.93  (0.84-1.04) | 0.98  (0.84-1.15) |  | 1.03  (0.96-1.11) | 1.10  (0.99-1.22) | 0.90  (0.76-1.06) |
| 5 | 0.91  (0.84-0.99) | 0.82  (0.73-0.92) | 1.13  (0.95-1.34) |  | 0.93  (0.86-1.00) | 1.01  (0.90-1.12) | 0.94  (0.80-1.11) |  | 1.05  (0.97-1.13) | 1.15  (1.03-1.28) | 0.96  (0.81-1.12) |
| *P*_trend_ | 0.057 | *<0.001* | *0.128* |  | *0.195* | *0.792* | *0.869* |  | *0.165* | *0.002* | *0.396* |

Abbreviations: CI, confidence interval; HR, hazards ratio; ref, reference.

^a^ Medians of dietary omega-3 percentage in quintiles 1-5 are 1.86, 2.37, 2.77, 3.30, 4.63.

^b^ Medians of dietary omega-6 percentage in quintiles 1-5 are 11.26, 14.01, 16.11, 18.49, 22.57.

^c^ Medians of dietary omega ratio in quintiles 1-5 are 3.49, 5.00, 5.88, 6.75, 8.25.

^d^ From Cox proportional hazards model; adjusted for dietary omega-6 percentage (; continuous), age (years; continuous), sex (male, female), race (White, Black, Asian, Others), Townsend deprivation index (continuous), assessment centre, BMI (kg/m2; continuous), smoking status (never, previous, current), alcohol intake status (never, previous, current), physical activity (low, moderate, high), and comorbidities (yes, no).

^e^ From Cox proportional hazards model; adjusted for dietary omega-3 percentage (; continuous), age (years; continuous), sex (male, female), race (White, Black, Asian, Others), Townsend deprivation index (continuous), assessment centre, BMI (kg/m2; continuous), smoking status (never, previous, current), alcohol intake status (never, previous, current), physical activity (low, moderate, high), and comorbidities (yes, no).

^f^ From Cox proportional hazards model; adjusted for age (years; continuous), sex (male, female), race (White, Black, Asian, Others), Townsend deprivation index (continuous), assessment centre, BMI (kg/m2; continuous), smoking status (never, previous, current), alcohol intake status (never, previous, current), physical activity (low, moderate, high), and comorbidities (yes, no).

**Table S7.** Selected participants serum biochemical markers at baseline across quintiles of the plasma omega-6/omega-3 PUFAs ratio (n=85,425).

|  | **Omega-6/omega-3 ratio quintiles** | | | | |  |
| --- | --- | --- | --- | --- | --- | --- |
| **Biomarker**^a^ | **1** (median = 5.9)  (*n* = 17,085) | **2** (median = 7.6)  (*n* = 17,085) | **3** (median = 9.1)  (*n* = 17,085) | **4** (median = 11.0)  (*n* = 17,085) | **5** (median = 14.8)  (*n* = 17,085) | *P* |
| **Cardiovascular:** |  |  |  |  |  |  |
| CRP (mg/L) | 2.3 (3.8) | 2.5 (4.2) | 2.5 (4.2) | 2.5 (4.2) | 2.7 (4.9) | <0.001^b^ |
| *Missing (n)* | *796* | *775* | *803* | *837* | *809* |  |
| **Cancer:** |  |  |  |  |  |  |
| SHBG (nmol/L) | 53.5 (30.0) | 51.8 (28.8) | 51.3 (27.7) | 51.4 (26.9) | 52.3 (26.2) | <0.001^b^ |
| *Missing (n)* | *2,280* | *2,229* | *2,364* | *2,312* | *2,231* |  |
| TTST (nmol/L) | 5.8 (5.7) | 6.3 (5.8) | 6.7 (6.0) | 7.1 (6.3) | 7.8 (6.5) | <0.001^b^ |
| *Missing (n)* | *2,806* | *2,461* | *2,187* | *2,081* | *1,819* |  |
| E2 (pmol/L) | 445.2 (383.7) | 470.2 (425.8) | 473.9 (503.8) | 457.0 (376.9) | 462.3 (382.1) | <0.001^b^ |
| *Missing (n)* | *14,910* | *14,777* | *14,174* | *13,958* | *13,382* |  |
| IGF-1 (nmol/L) | 21.5 (5.6) | 21.5 (5.5) | 21.7 (5.6) | 21.6 (5.8) | 21.4 (5.9) | <0.001^b^ |
| *Missing (n)* | *864* | *847* | *868* | *886* | *865* |  |

Abbreviations: CRP, C-creative protein; SHBG, sex hormone binding globulin; TTST, testosterone; E2, oestradiol; IGF-1, insulin-like growth factor 1.

^a^ All variables measured at baseline are presented as mean (SD) unless otherwise specified.

^b^ From the ANOVA test for continuous variables.

**Table S8.** Mediation analysis of biomarkers on associations between plasma omega-6/omega-3 ratio and mortality.

|  |  | Path a^a^  (Association between omega-6/omega-3 ratio and biomarker) | Path b^b^  (Association between biomarker and mortality) | Path c^b^  (Association between omega-6/omega-3 ratio and mortality) | Indirect effect | Proportion mediated |
| --- | --- | --- | --- | --- | --- | --- |
| Mortality | Biomarker | Coefficient  (95% CI) | HR Estimate  (95% CI) | HR Estimate  (95% CI) | Estimate  (95% CI) |  |
| All-cause | **Cardiovascular-related:** | | | | | |
|  | CRP | 0.037  (0.029-0.045) | 1.021  (1.016-1.025) | 1.020  (1.014-1.025) | 1.001  (1.001-1.001) | 4.0% |
|  | **Cancer-related:** | | | | | |
|  | SHBG | 0.179  (0.134-0.224) | 1.005  (1.004-1.006) | 1.019  (1.013-1.025) | 1.001  (1.001-1.001) | 5.0% |
|  | TTST | 0.057  (0.051-0.063) | 0.999  (0.989-1.009) | 1.020  (1.015-1.026) | 1.000  (0.999-1.000) | NA |
|  | E2 | -0.066  (-1.565-1.433) | 1.000  (1.000-1.000) | 1.027  (1.013-1.040) | 1.000  (1.000-1.000) | NA |
|  | IGF-1 | -0.084  (-0.094 - -0.074) | 0.992  (0.987-0.998) | 1.020  (1.014-1.025) | 1.001  (1.000-1.001) | 3.4% |
| Cardiovascular | CRP | 0.037  (0.029-0.045) | 1.012  (1.002-1.022) | 1.021  (1.010-1.031) | 1.000  (1.000-1.001) | 2.1% |
| Cancer | SHBG | 0.179  (0.134-0.224) | 1.003  (1.001-1.005) | 1.009  (0.999-1.019) | 1.000  (1.000-1.001) | 5.2% |
|  | TTST | 0.057  (0.051-0.063) | 0.980  (0.965-0.996) | 1.011  (1.001-1.021) | 0.999  (0.998-0.999) | NA |
|  | E2 | -0.066  (-1.565-1.433) | 1.000  (1.000-1.001) | 1.030  (1.009-1.052) | 1.000  (1.000-1.000) | NA |
|  | IGF-1 | -0.084  (-0.094 - -0.074) | 0.999  (0.991-1.007) | 1.012  (1.002-1.022) | 1.000  (0.999-1.000) | 0.8% |

Abbreviations: CRP, C-creative protein; SHBG, sex hormone binding globulin; TTST, testosterone; E2, oestradiol; IGF-1, insulin-like growth factor 1.

^a^ From linear regression model; age (years; continuous), sex (male, female), race (White, Black, Asian, Others), Townsend deprivation index (continuous), assessment centre, BMI (kg/m2; continuous), smoking status (never, previous, current), alcohol intake status (never, previous, current), physical activity (low, moderate, high), and comorbidities (yes, no).

^b^ From Cox proportional hazards model; age (years; continuous), sex (male, female), race (White, Black, Asian, Others), Townsend deprivation index (continuous), assessment centre, BMI (kg/m2; continuous), smoking status (never, previous, current), alcohol intake status (never, previous, current), physical activity (low, moderate, high), and comorbidities (yes, no).

^c^ NA: proportion mediated was not calculated when the point estimate of the direct effect was in an opposite direction to that of the indirect effect.

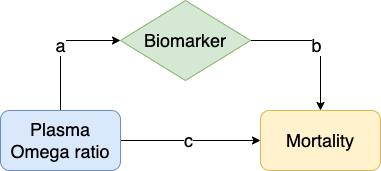

**Figure S1.** **Directed acyclic graph to explain mediation.** The ab arrows represent the indirect effect (i.e., the pathway through the mediator), while the c arrow represents the direct effects (i.e., all pathways other than through the mediator). Sum them up get the total effect which is the overall effect of exposure on outcome in the presence of a mediator.

**Table S9.** Associations^a^ of DHA and LA with all-cause, cancer, and CVD mortality risk in the UK Biobank.

|  | **DHA%**^c^ | | |  | **LA%**^d^ | | |
| --- | --- | --- | --- | --- | --- | --- | --- |
| **Cause of death** | All-cause | Cancer | CVD |  | All-cause | Cancer | CVD |
|  | HR  (95% CI) | HR  (95% CI) | HR  (95% CI) |  | HR  (95% CI) | HR  (95% CI) | HR  (95% CI) |
| Continuous | 0.91  (0.87-0.95) | 0.92  (0.86-0.98) | 0.91  (0.84-1.00) |  | 0.96  (0.96-0.97) | 0.97  (0.95-0.98) | 0.97  (0.96-0.99) |
| Quintiles^a,b^ |  |  |  |  |  |  |  |
| 1 | 1.00 (ref) | 1.00 (ref) | 1.00 (ref) |  | 1.00 (ref) | 1.00 (ref) | 1.00 (ref) |
| 2 | 0.96  (0.88-1.04) | 1.06  (0.93-1.21) | 0.89  (0.76-1.05) |  | 0.86  (0.79-0.93) | 0.86  (0.76-0.97) | 0.84  (0.72-0.98) |
| 3 | 0.88  (0.80-0.96) | 0.98  (0.85-1.12) | 0.75  (0.62-0.89) |  | 0.81  (0.74-0.88) | 0.81  (0.71-0.92) | 0.87  (0.74-1.03) |
| 4 | 0.83  (0.76-0.91) | 0.90  (0.79-1.04) | 0.81  (0.68-0.97) |  | 0.80  (0.73-0.88) | 0.77  (0.67-0.88) | 0.90  (0.75-1.08) |
| 5 | 0.81  (0.74-0.89) | 0.85  (0.74-0.98) | 0.77  (0.64-0.93) |  | 0.66  (0.60-0.73) | 0.69  (0.60-0.81) | 0.67  (0.54-0.83) |
| *P*_trend_ | *<0.001* | *0.003* | *0.003* |  | *<0.001* | *<0.001* | *0.001* |

Abbreviations: DHA%, docosahexaenoic acid to total fatty acids percentage; LA%, linoleic acid to total fatty acids percentage; CI, confidence interval; HR, hazards ratio; ref, reference.

^a^ Medians of DHA percentage in quintiles 1-5 are 1.24, 1.64, 1.92, 2.25, 2.84.

^b^ Medians of LA percentage in quintiles 1-5 are 24.8, 27.6, 29.4, 31.1, 33.4.

^c^ Adjusted for LA percentage, age (years; continuous), sex (male, female), race (White, Black, Asian, Others), Townsend deprivation index (continuous), assessment centre, BMI (kg/m2; continuous), smoking status (never, previous, current), alcohol intake status (never, previous, current), physical activity (low, moderate, high), and comorbidities (yes, no).

^d^ Adjusted for DHA percentage, age (years; continuous), sex (male, female), race (White, Black, Asian, Others), Townsend deprivation index (continuous), assessment centre, BMI (kg/m2; continuous), smoking status (never, previous, current), alcohol intake status (never, previous, current), physical activity (low, moderate, high), and comorbidities (yes, no).

**Table S10.** Associations^a^ of ratio of omega-6/omega-3 PUFAs with all-cause, cancer, and CVD mortality risk in the UK Biobank, covariates including fish oil supplementation status.

| **Omega ratio variable forms** | **Causes of death** | | | | | | | | | | | |
| --- | --- | --- | --- | --- | --- | --- | --- | --- | --- | --- | --- | --- |
|  | **All-cause** | | |  | **Cancer** | | |  | **Cardiovascular diseases** | | | |
|  | Number of deaths | Partially adjusted associations^b^ | Fully adjusted associations^c^ |  | Number of deaths | Partially adjusted associations^b^ | Fully adjusted associations^c^ |  | Number of deaths | Partially adjusted associations^b^ | | Fully adjusted associations^c^ |
|  |  | HR  (95% CI) | HR  (95% CI) |  |  | HR  (95% CI) | HR  (95% CI) |  |  | HR  (95% CI) | HR  (95% CI) | |
| Continuous | 6,461 | 1.02  (1.02-1.03) | 1.02  (1.01-1.03) |  | 2,794 | 1.02  (1.01-1.03) | 1.01  (1.00-1.02) |  | 1,668 | 1.01  (1.00-1.02) | 1.02  (1.01-1.03) | |
| Quintiles  (median) |  |  |  |  |  |  |  |  |  |  |  | |
| 1 (5.9) | 1,348 | 1.00 (ref) | 1.00 (ref) |  | 593 | 1.00 (ref) | 1.00 (ref) |  | 369 | 1.00 (ref) | 1.00 (ref) | |
| 2 (7.6) | 1,256 | 0.99  (0.91-1.06) | 0.95  (0.87-1.04) |  | 563 | 1.01  (0.90-1.14) | 0.97  (0.85-1.11) |  | 315 | 0.89  (0.76-1.03) | 0.88  (0.74-1.05) | |
| 3 (9.1) | 1,236 | 1.04  (0.96-1.12) | 1.00  (0.91-1.09) |  | 543 | 1.06  (0.94-1.19) | 0.98  (0.85-1.12) |  | 321 | 0.95  (0.81-1.10) | 0.96  (0.80-1.15) | |
| 4 (11.0) | 1,252 | 1.11  (1.03-1.20) | 1.06  (0.97-1.17) |  | 548 | 1.14  (1.01-1.29) | 1.09  (0.95-1.25) |  | 306 | 0.95  (0.81-1.11) | 1.00  (0.84-1.20) | |
| 5 (14.8) | 1,369 | 1.30  (1.20-1.40) | 1.23  (1.12-1.34) |  | 547 | 1.23  (1.09-1.39) | 1.11  (0.97-1.28) |  | 357 | 1.15  (0.99-1.34) | 1.27  (1.07-1.52) | |
| *P*_trend_ |  | *<0.001* | *<0.001* |  |  | *<0.001* | *0.037* |  |  | *0.018* | *<0.001* | |

Abbreviations: CI, confidence interval; HR, hazards ratio; ref, reference.

^a^ From Cox proportional hazards regression.

^b^ Adjusted for age (years; continuous), sex (male, female), race (White, Black, Asian, Others), Townsend deprivation index (continuous), assessment centre, fish oil supplementation.

^c^ Adjusted for age (years; continuous), sex (male, female), race (White, Black, Asian, Others), Townsend deprivation index (continuous), assessment centre, BMI (kg/m2; continuous), smoking status (never, previous, current), alcohol intake status (never, previous, current), physical activity (low, moderate, high), comorbidities (yes, no), and fish oil supplementation.

**Table S11.** Associations^a^ of ratio of omega-6/omega-3 PUFAs with all-cause, cancer, and CVD mortality risk in the UK Biobank Study, with multiple imputation for missing data.

| **Omega ratio variable forms** | **Causes of death** | | | | | | | | | | | |
| --- | --- | --- | --- | --- | --- | --- | --- | --- | --- | --- | --- | --- |
|  | **All-cause** | | |  | **Cancer** | | |  | **Cardiovascular diseases** | | | |
|  | Number of deaths | Partially adjusted associations^b^ | Fully adjusted associations^c^ |  | Number of deaths | Partially adjusted associations^b^ | Fully adjusted associations^c^ |  | Number of deaths | Partially adjusted associations^b^ | | Fully adjusted associations^c^ |
|  |  | HR  (95% CI) | HR  (95% CI) |  |  | HR  (95% CI) | HR  (95% CI) |  |  | HR  (95% CI) | HR  (95% CI) | |
| Continuous | 6,461 | 1.02  (1.02-1.03) | 1.02  (1.01-1.02) |  | 2,794 | 1.02  (1.01-1.03) | 1.02  (1.01-1.02) |  | 1,668 | 1.02  (1.01-1.03) | 1.01  (1.00-1.02) | |
| Quintiles  (median) |  |  |  |  |  |  |  |  |  |  |  | |
| 1 (5.9) | 1,348 | 1.00 (ref) | 1.00 (ref) |  | 593 | 1.00 (ref) | 1.00 (ref) |  | 369 | 1.00 (ref) | 1.00 (ref) | |
| 2 (7.6) | 1,256 | 0.99  (0.92-1.07) | 0.97  (0.90-1.05) |  | 563 | 1.02  (0.91-1.15) | 1.00  (0.89-1.12) |  | 315 | 0.89  (0.77-1.04) | 0.87  (0.74-1.01) | |
| 3 (9.1) | 1,236 | 1.06  (0.98-1.15) | 1.05  (0.97-1.13) |  | 543 | 1.08  (0.96-1.21) | 1.05  (0.94-1.19) |  | 321 | 0.97  (0.84-1.13) | 0.97  (0.83-1.12) | |
| 4 (11.0) | 1,252 | 1.14  (1.05-1.23) | 1.10  (1.02-1.19) |  | 548 | 1.16  (1.03-1.30) | 1.12  (1.00-1.26) |  | 306 | 0.97  (0.84-1.13) | 0.95  (0.82-1.11) | |
| 5 (14.8) | 1,369 | 1.34  (1.24-1.44) | 1.28  (1.19-1.38) |  | 547 | 1.26  (1.12-1.42) | 1.20  (1.07-1.35) |  | 357 | 1.19  (1.03-1.38) | 1.17  (1.01-1.36) | |
| *P*_trend_ |  | *<0.001* | *<0.001* |  |  | *<0.001* | *<0.001* |  |  | *0.004* | *<0.001* | |

Abbreviations: CI, confidence interval; HR, hazards ratio; ref, reference.

^a^ From Cox proportional hazards regression.

^b^ Adjusted for age (years; continuous), sex (male, female), race (White, Black, Asian, Others), Townsend deprivation index (continuous), assessment centre.

^c^ Adjusted for age (years; continuous), sex (male, female), race (White, Black, Asian, Others), Townsend deprivation index (continuous), assessment centre, BMI (kg/m2; continuous), smoking status (never, previous, current), alcohol intake status (never, previous, current), physical activity (low, moderate, high), and comorbidities (yes, no).

**Table S12.** Associations^a^ of ratio of omega-6/omega-3 PUFAs with all-cause, cancer, and CVD mortality risk in the UK Biobank Study, excluding those who died in the first follow-up year.

| **Omega ratio variable forms** | **Causes of death** | | | | | | | | | | | |
| --- | --- | --- | --- | --- | --- | --- | --- | --- | --- | --- | --- | --- |
|  | **All-cause** | | |  | **Cancer** | | |  | **Cardiovascular diseases** | | | |
|  | Number of deaths | Partially adjusted associations^b^ | Fully adjusted associations^c^ |  | Number of deaths | Partially adjusted associations^b^ | Fully adjusted associations^c^ |  | Number of deaths | Partially adjusted associations^b^ | | Fully adjusted associations^c^ |
|  |  | HR  (95% CI) | HR  (95% CI) |  |  | HR  (95% CI) | HR  (95% CI) |  |  | HR  (95% CI) | HR  (95% CI) | |
| Continuous | 6,345 | 1.02  (1.02-1.03) | 1.02  (1.02-1.03) |  | 2,754 | 1.02  (1.01-1.03) | 1.01  (1.00-1.02) |  | 1,620 | 1.02  (1.01-1.03) | 1.02  (1.01-1.03) | |
| Quintiles  (median) |  |  |  |  |  |  |  |  |  |  |  | |
| 1 (5.9) | 1,348 | 1.00 (ref) | 1.00 (ref) |  | 588 | 1.00 (ref) | 1.00 (ref) |  | 360 | 1.00 (ref) | 1.00 (ref) | |
| 2 (7.6) | 1,235 | 1.00  (0.92-1.08) | 0.96  (0.88-1.05) |  | 555 | 1.02  (0.91-1.15) | 0.97  (0.85-1.11) |  | 306 | 0.90  (0.77-1.04) | 0.89  (0.75-1.07) | |
| 3 (9.1) | 1,217 | 1.06  (0.98-1.15) | 1.02  (0.93-1.12) |  | 538 | 1.08  (0.96-1.21) | 1.00  (0.87-1.14) |  | 314 | 0.98  (0.84-1.14) | 0.98  (0.82-1.18) | |
| 4 (11.0) | 1,225 | 1.13  (1.05-1.23) | 1.08  (0.99-1.19) |  | 537 | 1.15  (1.02-1.29) | 1.10  (0.97-1.26) |  | 296 | 0.97  (0.83-1.14) | 1.02  (0.85-1.22) | |
| 5 (14.8) | 1,340 | 1.34  (1.24-1.44) | 1.26  (1.15-1.37) |  | 536 | 1.25  (1.11-1.41) | 1.14  (0.99-1.30) |  | 344 | 1.19  (1.02-1.38) | 1.29  (1.08-1.53) | |
| *P*_trend_ |  | *<0.001* | *<0.001* |  |  | *<0.001* | *0.016* |  |  | *0.005* | *<0.001* | |

Abbreviations: CI, confidence interval; HR, hazards ratio; ref, reference.

^a^ From Cox proportional hazards regression.

^b^ Adjusted for age (years; continuous), sex (male, female), race (White, Black, Asian, Others), Townsend deprivation index (continuous), assessment centre.

^c^ Adjusted for age (years; continuous), sex (male, female), race (White, Black, Asian, Others), Townsend deprivation index (continuous), assessment centre, BMI (kg/m2; continuous), smoking status (never, previous, current), alcohol intake status (never, previous, current), physical activity (low, moderate, high), and comorbidities (yes, no).

**Table S13**. Baseline characteristics of participants with missing exposure information and not included in the study.

| Characteristics^a^ | Not included  (N=416,959) | Included  (N=85,425) |
| --- | --- | --- |
| Age (years) | 56.7 (8.1) | 55.9 (8.2) |
| Gender (male%) | 45.3 | 47.0 |
| Ethnicity(n%) |  |  |
| White | 376,904 (90.9%) | 77,242 (90.9%) |
| Black | 2,347 (0.6%) | 525 (0.6%) |
| Asian | 15,842 (3.8%) | 3,287 (3.9%) |
| Others | 19,504 (4.7%) | 3,955 (4.7%) |
| *Missing (n)* | *2,362* | *416* |
| TDI | -1.3 (3.1) | -1.3 (3.1) |
| *Missing (n)* | *511* | *115* |
| BMI | 27.5 (4.8) | 27.2 (4.7) |
| *Missing (n)* | *2,805* | *302* |
| Smoking status (n%) |  |  |
| Never | 226,722 (54.7%) | 46,736 (55.0%) |
| Previous | 144,202 (34.8%) | 28,811 (33.9%) |
| Current | 43,536 (10.5%) | 9,426 (11.1%) |
| *Missing (n)* | *2499* | *452* |
| Alcohol status (n%) |  |  |
| Never | 18,689 (4.5%) | 3,691 (4.3%) |
| Previous | 15,016 (3.6%) | 3,077 (3.6%) |
| Current | 381,825 (91.9%) | 78,431 (92.1%) |
| *Missing (n)* | *1429* | *226* |
| Physical activity (n%) |  |  |
| Low | 63,283 (19.0%) | 12,907 (18.6%) |
| Moderate | 135,778 (40.8%) | 28,213 (40.7%) |
| High | 133,905 (40.2%) | 28,194 (40.7%) |
| *Missing (n)* | *83,993* | *16,111* |
| Fish oil supplementation (Yes%) |  |  |
|  | 31.5 | 30.7 |
| *Missing (n)* | *5,869* | *327* |

^a^ All variables measured at baseline are presented as mean (SD) except as otherwise specified.
